# TabMedQA: From Structured Data to Question-Answer Datasets in Early Clinical Decision-Making

**DOI:** 10.64898/2026.08.19.26360779

**Authors:** Gabriel Iturra-Bocaz, Petra Galuščáková, Sol Vedde, Alvaro Fernandez-Quilez

**Affiliations:** University of Stavanger; Stavanger University Hospital

**Keywords:** Medical QA Datasets, Tabular Data, EHR, Retrieval Augmented Generation, decision-making

## Abstract

The rising adoption of Large Language Models (LLMs) and Retrieval Augmented Generation (RAG) in clinical general practice demands datasets that capture realistic early-stage clinical decision-making, where experts must decide on follow-up actions based on sparse, structured patient data. Existing medical Question–Answering (QA) resources primarily address post-diagnostic or specialist settings and rarely reflect how General Practitioners (GPs) document and justify early decisions based on clinical observations from Electronic Health Records (EHRs) and grounded on clinical guidelines. We present TabMedQA, a framework for synthesizing QA collections that emulate how GPs formulate and document decisions in encounter notes during early patient assessments. TabMedQA leverages instruction-tuned LLMs, guided by disease-specific clinical guidelines, to generate full encounter notes composed of a guideline-grounded justification and a corresponding follow-up recommendation directly from structured EHR inputs. The framework further supports RAG-based evaluation, simulating how GPs might consult previous patient encounters to inform new consultations. We demonstrate the application and resulting resource use of TabMedQA on prostate cancer using the publicly available PI-CAI collection and release the resulting PI-CAI QA collection, resource generation templates, and TabMedQA code. To the best of our knowledge, TabMedQA provides the first open framework for creating guideline-grounded, EHR-based QA collections that enable the generation and holistic evaluation of LLM-produced clinical encounter notes, bridging decision-making accuracy with clinical encounter quality in general practice.

## 1. Introduction

General Practitioners (GPs) operate at the earliest stage of clinical decision-making, where they have to interpret sparse and highly heterogeneous patient information to propose a clinically grounded and justified course of action (Starfield et al., 2005). For this purpose, they typically rely on limited structured data from Electronic Health Records (EHRs) such as demographics, risk factors and medical test screening results (Crosson et al., 2005). By the end of each patient consultation, the GP must document a medical encounter note that includes (i) a recommendation regarding patient follow-up and (ii) a justification of their decision grounded in clinical observations and established medical guidelines (Europe, 2002). These encounter notes serve not only as formal documentation of the consultation, but also as a legal and ethical safeguard ensuring traceability of decisions and transparency towards patients (Rose and Joshi, 2018; Sittig and Singh, 2011).

Large Language Models (LLMs) equipped with Retrieval Augmented Generation (RAG) are increasingly integrated into clinical workflows of GPs to assist with documentation, information retrieval and decision support (Gaber et al., 2025; Lu et al., 2024). Such systems can access collections of previous patient encounters and their medical notes,allowing them to ground their responses in comparable cases. Within this context, the process of generating a medical encounter note from EHR data can be naturally viewed as a question-answering (QA) task (Figure 1). Here, the GP acts as the user who formulates a question to an LLM system that has access to the patient’s structured information, and, optionally, similar historical cases, requesting an answer in the form of an encounter note that outlines an appropriate follow-up recommendation together with a justification consistent with clinical guidelines. However, in spite of the relevance of the scenario in real-world clinical practice, collections supporting the systematic development and evaluation of LLM with RAG emulating GP-LLM interaction to produce clinical encounter notes remain scarce (Shool et al., 2025; Elgedawy et al., 2024; Gomez-Cabello et al., 2024).

**Figure 1:**
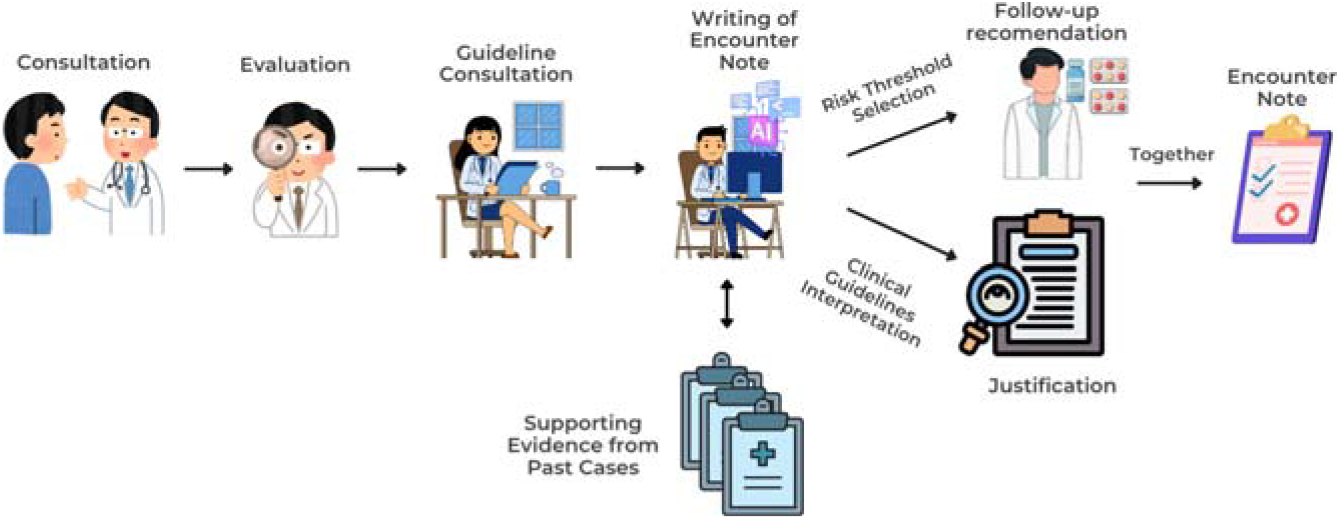
Overview of a GP’s consultation workflow, from patient evaluation to the creation of an encounter note containing a follow-up recommendation and its clinical justification.

Existing clinical QA datasets primarily address specialist or post-diagnostic settings, relying on the availability of medical notes written by experts or extracting answers from complete clinical narratives (Jin et al., 2021; Singhal et al., 2025; Kim et al., 2024). In both cases, these resources are difficult to obtain at scale because of privacy restrictions, the high cost of expert annotation and the variability in clinical documentation practices (Williamson and Prybutok, 2024; Kitamura et al.,2024; Cohen et al., 2019). Furthermore, existing clinical QA collections provide brief factual answers or diagnostic labels rather than guideline-grounded encounter notes that explicitly articulate both the justification behind a clinical decision and the corresponding decision (Kim et al., 2024). As a result, current datasets and benchmarks provide limited support for studying how can LLMs generate guideline-consistent encounter notes from EHR structured data, or how to perform realistic and holistic evaluations of encounter notes including decision-making and overall clinical encounter note quality (Bardhan et al., 2024).

To address these gaps, we introduce **TabMedQA**, a framework to construct medical QA collections that simulate how GPs could interact with RAG-equipped LLMs during initial patient evaluations to produce medical encounter notes. The framework leverages instruction-tuned LLMs guided by disease-specific clinical guidelines to generate question-answer pairs directly from structured EHRs, where each answer corresponds to an encounter note combining a clinical-guideline grounded justification and a follow-up recommendation (Figure 2).

**Figure 2:**
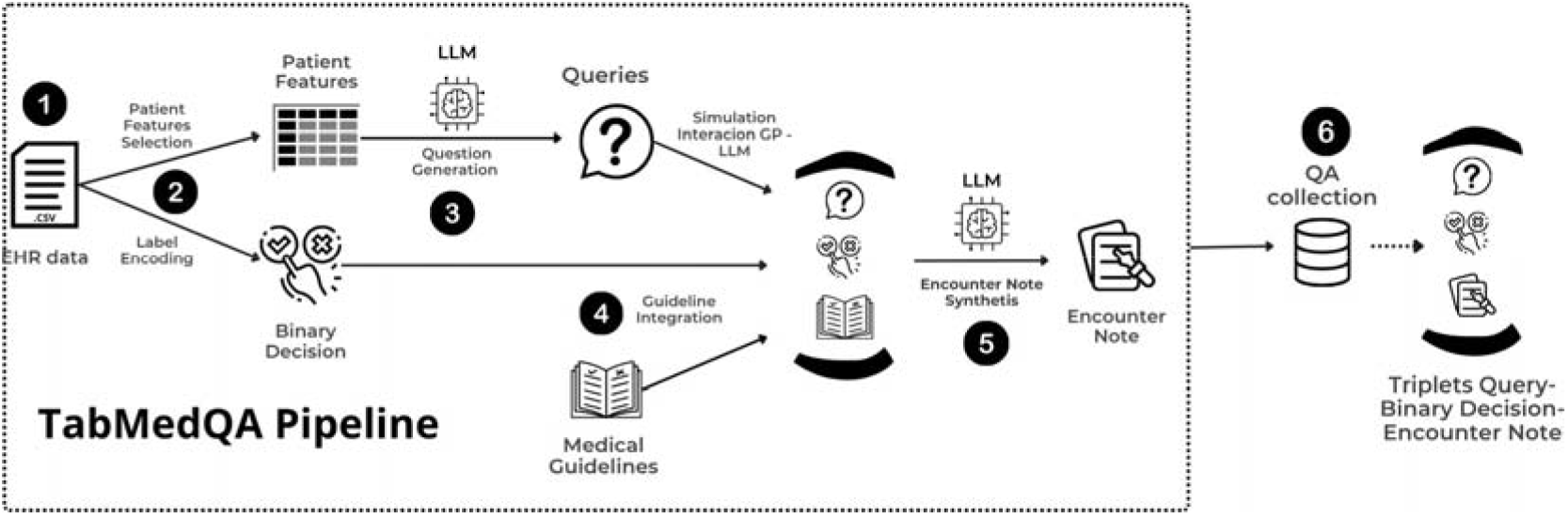
Overview of the **TabMedQA** pipeline: From structured (1) EHR data, (2) *EHR Preprocessing* (patient feature selection and label encoding), (3) *Question Generation*, (4–5) *Guideline Integration, Encounter Note Synthesis*, and (6) the resulting *QA collection*.

At inference time, resources generated with TabMedQA support RAG-based evaluation, simulating how a GP might consult an automated system that retrieves previous patient encounter notes to inform new follow-up recommendations and justifications (Figure 1). We demonstrate its use through one study case to depict the application of TabMedQA framework and resulting QA collection in a clinically realistic early decision-making scenario. Our main contributions can be summarized as follows:

- We introduce **TabMedQA**, a framework for constructing medical QA collections that simulate early general-practice consultations (Figure 1). TabMedQA synthesizes encounter notes with both clinical justifications and follow-up recommendations from structured EHR data, guided by disease-specific clinical guidelines and instructed LLMs.
- We show how TabMedQA-generated QA collections can be used in clinical real-world scenarios in which an LLM accesses **a bank of previous patient cases** to inform recommendations and justifications for a given new patient EHR and visit. This mirrors how GPs could interact with LLM-based systems in clinical practice during new patient visits.
- We release a QA collection focuses on prostate cancer (PI-CAI QA), together with the corresponding generation templates and TabMedQA code to support future research on early-stage justified clinical-decision making^1^.

The remainder of the paper is organized as follows: Section 2 reviews relevant literature, Section 3 presents the methodological approach to construct the **TabMedQA** framework, Section 4 describes the resulting QA collection obtained by applying TabMedQA to a clinical use case and provides guidance on how the resource can be leveraged at inference time to simulate GP–LLM interactions, Following, Section 5 details experiments evaluating these interactions in RAG-based settings where prior encounter notes are used as a support to synthesize new encounter notes. Section 6 discusses results and findings, whilst Section 7 concludes with perspectives for future work. Finally, Section 8 we discuss the limitation our proposed TabMedQA framework.

## 2. Related Work

The rapid adoption and integration of LLMs equipped with RAG in clinical practice has increased the research community interest in under-standing their impact in decision support and automatic documentation (Eguia et al., 2024; Gaber et al., 2025). However, most existing work in this area has focused on specialist or post-diagnostic clinical scenarios (Ke et al., 2025), with limited attention to how they could support early, GP settings where data are primarily structured and decisions must be justified according to clinical guidelines (Gomez-Cabello et al., 2024; Korom et al., 2025). In this section, we review related work along two complementary directions: (i) the use of RAG systems in healthcare applications and (ii) the development of QA datasets for evaluating clinical LLMs. Together, they highlight the absence of resources that combine structured EHR data, guideline-grounded justifications and retrieval-based access to prior patient cases emulating realistic GP workflows (Figure 1).

### 2.1. RAG in Healthcare

In the healthcare domain, RAG systems have been explored for a variety of applications, including the interpretation of clinical guidelines, clinical decision-support, and diagnostic assistance (Amugongo et al., 2025; Ge et al., 2024; Rau et al., 2024). For example, Ge et al. (2024) introduced LiVersa, a liver disease-specific chat bot that leverages RAG systems to perform clinical answering for hepatology questions. Similarly, Rau et al. (2024) developed a RAG-based chat bot designed for gastrointestinal imaging, which achieved significantly higher accuracy than generic GPT-4 models. Recent studies have also applied RAG to medical QA and literature-grounded reasoning tasks, retrieving evidence from textual biomedical sources rather than patient data (Singhal et al., 2025; Zhang et al., 2025). While these systems demonstrate the potential of RAG in specialized clinical contexts, they primarily rely on retrieving textual medical knowledge to support post-diagnostic decision-making. In contrast, the use of RAG for patient-case retrieval, where models draw on previous clinical encounters notes or EHR-derived cases, remains largely unexplored. This gap is particularly evident in the pre-diagnostic stage of GPs, where GPs must interpret sparse structured data to justify and document initial decisions.

### 2.2. QA Collections for Early Clinical Decision-making

Benchmark QA collections are essential for evaluating LLMs and RAG systems, particularly in domain-specific settings such as healthcare. Existing collections, such as MedQA, MedMCQA, and PubMedQA (Jin et al., 2021; Pal et al., 2022; Jin et al., 2019) typically consist of multiple-choice questions or short patient histories designed to assess models’ abilities to answer clinical questions. However, these datasets generally provide only question-response pairs, without capturing the guideline-grounded justifications that clinicians are expected to provide when formulating or documenting medical decisions. In real-world practice, especially in GP settings, transparency regarding the rationale and clinical guideline alignment behind a decision is necessary for accountability and communication with patients (Kim et al., 2024).

In an effort to move towards this direction, Kim et al. (2024) introduced MedExQA, which augments QA pairs with manually crafted explanations describing the clinical rationale behind each answer. Another example is PMC-Patients (Zhao et al., 2023), a large patient-summary retrieval dataset designed for retrieval-based clinical decision support, which supports patient-to-patient and patient-to-article retrieval tasks. This setup is conceptually similar to our approach of retrieving clinically similar patient cases to inform new decisions. While MedExQA and PMC-Patients represent important steps, their focus remains on specialist or post-diagnostic scenarios, and its explanations are generated through manual supervision rather than structured EHR data and formalized clinical guidelines.

## 3. TabMedQA Framework Methodology

In TabMedQA, the process of constructing question–medical encounter note pairs (QA), follows a systematic pipeline consisting of four main components: EHR Pre-processing, Question Generation, Guideline Integration, and Encounter Note Synthesis. We illustrate the overall workflow in Figure 2. To demonstrate the applicability of the framework, we apply TabMedQA to a use case focuses on prostate cancer (Saha et al. (2024)), which is described in the following subsections.

### 3.1. Prostate Imaging-Cancer Collection

Our framework is applied to the Prostate Imaging-Cancer collection (PI-CAI) introduced by Saha et al. (2024). PI-CAI^2^ is a multi-modal benchmark for evaluating the detection of clinically significant prostate cancer (csPC) in a specialist diagnostic setting. The original collection^3^ comprises 1,500 MRI examinations from 1,476 male patients clinically suspected of having csPC and referred for specialist testing between 2012 and 2021. The data were collected from several hospitals in the Netherlands (Saha et al., 2024; Gade et al., 2024). Each patient case includes both imaging data and an EHR^4^ file describing clinical and demographic patient features together with a expert-verified label indicating whether follow-up was necessary under the suspicion of csPC presence (Fernandez-Quilez et al., 2024). This decision label is leveraged as the patient follow-up recommendation ground truth in our pipeline (Figure 2).

For the purpose of our work, we apply the following inclusion criteria to the original available collection (i) EHR records with no missing values for prostate specific antigen (PSA), prostate specific antigen density (PSAd), prostate volume, patient age or follow-up decision ground truth (ii) availability of all these features at patient’s first recorded consultation. After applying these criteria, the resulting collection use for our study case comprised 606 patient EHR records from different patients (Figure 2, Step 1).

### 3.2. EHR Preprocessing

The EHR Pre-processing stage (Figure 2, Step 2) is designed to prepare structured patient features for TabMedQA by retaining only the features that mirror the type of information GPs access during early diagnostic evaluations in primary care. Therefore, the following features are retained: patient age, PSA, PSAd, and prostate volume. The identification of these variables is conducted in collaboration with an expert and in alignment with clinical guidelines (Cornford et al., 2025).

The original PI-CAI decision label indicating whether each case is at risk of harboring csPC and thus requires follow-up, is simplified into a binary decision representing whether a patient is at risk (1) or not at risk (0) of harboring csPC.

### 3.3. Question Generation

The Question Generation stage (Figure 2, Step 3) aims to emulate the interaction between a GP and an LLM during an early-stage consultation, where the GP questions the system about the likelihood of a condition and its justification based on the available patient features. We choose to focus on likelihood estimation rather than binary decision based on real clinical workflows where GPs interpret these likelihoods against guideline-based risk thresholds to decide whether follow-up is necessary (Cohen et al., 2019). Each question is grounded in the patient’s structured EHR data, which provide the patient features available at the time of the consultation.

To operationalize this process, we employ a pretrained LLM, `gpt4o_mini`^5^ to produce medically relevant questions align with the emulated GP-LLM interaction. Although the proposed methodology is model-agnostic and can be with any instructed-tuned LLM, `gpt4o_mini` is selected for its demonstrated ability to generate high-quality and contextually relevant clinical QA pairs (Lunardi et al., 2025; Keat et al., 2024; Rewthamrongsris et al., 2025).

The model is prompted with a set of structured EHR features and instructed to generate clinically coherent questions that a GP might pose to the system when evaluating a new patient and writing the respective encounter note. The Prompt 1 is designed to ensure that generated questions are (i)grounded in the provided patient features, (ii) phrased in natural language and (iii) medically plausible. All generated questions are subsequently reviewed by a clinical expert to verify their validity and relevance.

#### Prompt 1: Query Generation

You are a General Practitioner expert. Given the following patient data columns:

{patient_feature_descriptions}

transform the provided tabular patient features into a medical question intended for General Practitioners and generate a JSON with the following format:

`{“question”: <generated_question>}`

**EHR data:** {patient_feature_values}

**Your output:**

### 3.4. Guideline Integration and Encounter Note Synthesis

Once the question is generated, TabMedQA synthesizes complete medical encounter notes anchored in clinical guidelines, combining guideline-grounded justification with the corresponding follow-up recommendation (Figure 2, Steps 4–5). This process emulates how experts justify follow-up recommendations in real consultations.

To incorporate the guidelines, we first adapt them to the target use case in collaboration with a clinical expert (Figure 2, Step 4). This integration step serves as a connection between the structured input and the model’s ability to produce interpretable justifications, regardless of the disease or dataset being used.

For the PI-CAI study, we employ the European Association of Urology (EAU) diagnostic guidelines (Cornford et al., 2025) which define recommended threshold values for PSA and PSAd across age groups. Due to the full EAU guidelines extend beyond what GPs typically apply during early consultations, we derive a simplified version verify by an expert capturing the practical recommendations that guide GP decision-making^6^. Whilst we refer to the study case, the approach is disease-agnostic: when official clinical guidelines are available for a condition they can be incorporated directly into the framework.

The final encounter note is then constructed (Figure 2, Step 5) by combining three elements: (i) the original GP-style question, which provides contextual understanding of what must be justified; (ii) the relevant guideline, which constrains the justification to evidence-based thresholds and recommendations; and (iii) the patient features, which anchors the justification in individual clinical findings. We employ `gpt4o_mini` as the generation model, guided by the Prompt 2.

#### Prompt 2: Encounter Note Synthesis

You are a General Practitioner expert writing an encounter note for a patient. Use the provided patient features and binary decision to justify the outcome according to the guidelines:{guidelines}.

For the question “{question}”, where the decision answer is {binary_decision}. Here, (1) indicates that the patient is prone to developing the disease in the future; (0) otherwise.

Provide a brief justification for General Practitioners, referencing how the patient features in the question correspond with the guidelines and considerations in clinical decision-making. Do not mention the label itself or use definitive diagnostic statements, but consider that the label indicates whether the patient has clinically significant prostate cancer or not. Use formal clinical language and relevant medical context, and do not include any information beyond what is provided.

Return your response in the following JSON format only:

`{ “encounter_note”: “<justification according to patient features and guidelines>” }`

**Your output:**

The prompt results in a synthesized encounter note, consisting of a concise justification and a follow-up recommendation. These synthesized encounter notes represent the final output of TabMedQA, combining medical evidence-based justifications with follow-up recommendations to simulate a realistic GP documentation.

## 4. PI-CAI QA Collection

After applying TabMedQA to the PI-CAI collection, we obtain the **PI-CAI QA collection**. PI-CAI QA is composed of 606 patient records represented as triplets (***q, d, n***), corresponding to the question, a binary decision, and the encounter note, respectively (Figure 2, Step 6).

To support experimental evaluation, we randomly divide the collection into a training and a test set that preserved the original preprocessed PI-CAI class distribution (1: 49%, 0: 51%). The training set consists of 484 unique triplet samples (≈ 80%), and serves as a bank of past patient uses cases. The test set contains 122 unique triplet samples (≈ 20%). The question ***q*** in the test set serves as the retrieval input, the corresponding encounter note ***n*** and the binary decision ***d*** act as the target ground truth. Detailed statistics of PI-CAI QA are presented in Table 1, whilst Table 2 illustrates one randomly selected triplet from the generated PI-CAI QA collection.

**Table 1:** Descriptive statistics of the PI-CAI QA dataset.

| Statistic | Value | Description |
| --- | --- | --- |
| Number of patient sessions | 606 | Total generated QA entries. |
| Number of patient features | 4 | Age, PSA, PSA<sub>d</sub>, prostate volume. |
| Binary classes | 2 (0 / 1) | Non-prone vs. prone to clinically significant PCa. |
| Class distribution | 1 / 0 (49% / 51%) | Balanced label ratio. |
| Training / Test split | 484 / 122 (80% / 20%) | Stratified by decision label. |
| Avg. question length | 68.24 tokens | Mean number of tokens per question. |
| Avg. justification length | 165.1 tokens | Mean number of tokens per encounter note. |
| Total number of tokens | 141,229 | Including questions and encounter notes. |
| Total triplets | 606 | Number of (q, d, n) entries. |

**Table 2:**
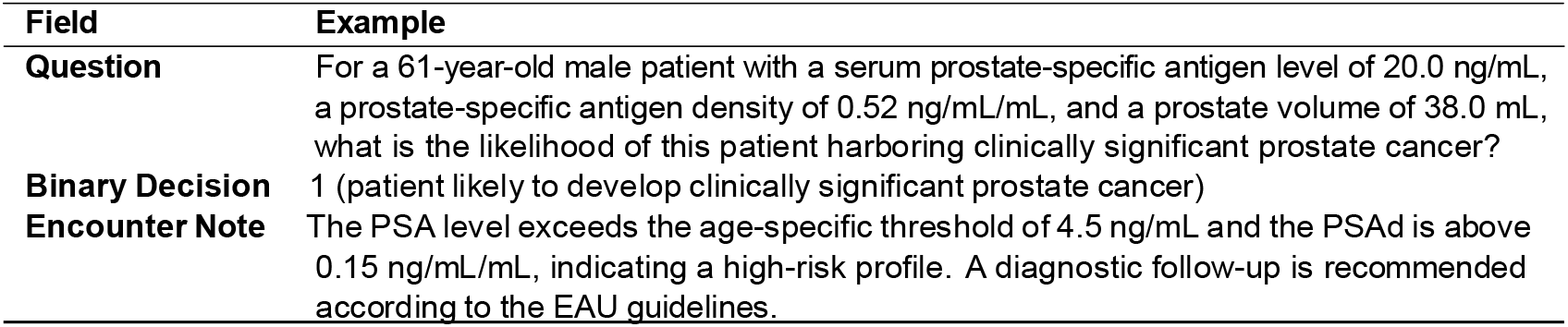
Example from the PI-CAI QA collection. Each triplet includes a question, the ground truth for the binary decision, and a guideline-grounded encounter note.

| Field | Example |
| --- | --- |
| **Question** | For a 61-year-old male patient with a serum prostate-specific antigen level of 20.0 ng/mL, a prostate-specific antigen density of 0.52 ng/mL/mL, and a prostate volume of 38.0 mL, what is the likelihood of this patient harboring clinically significant prostate cancer? |
| **Binary Decision** | 1 (patient likely to develop clinically significant prostate cancer) |
| **Encounter Note** | The PSA level exceeds the age-specific threshold of 4.5 ng/mL and the PSA<sub>d</sub> is above 0.15 ng/mL/mL, indicating a high-risk profile. A diagnostic follow-up is recommended according to the EAU guidelines. |

## 5. Experiments

To verify the usability of the collection, we apply several models to it. The experiments comprise two tasks that seek to reflect the dual goals of TabMedQA: (i) binary decision classification, and(ii)generation of encounter notes that simulate the ones a GP might write after a consultation including both the decision and its corresponding clinical guideline-grounded justification.

### Model Descriptions

We evaluate four models representing both feature-based and LLM-based approaches:

- **KNN:** A-nearest neighbors classifier trained on structured EHR data including patients’ age, PSA, PSAd and prostate volume. Each test case is labeled according to the majority class among k most similar training patient, computed via Minkowski distance over structured patient features. The optimal number of neighbors (k = 19) is selected through grid search using 10-fold cross validation.
- **KNN leveraging Encounter Note Embeddings:** Encounters notes from the training set and test questions are embedded using Clinical ModernBERT^7^ (Lee et al., 2025). We select this model due to its strong performance on clinical text embeddings, and for ensuring a fair comparison with the RAG that also relies on the same model. The classification task is performed by comparing each test question to the embedded encounter notes in this shared vector space. The best configuration is ***k*** = 41, also determined via grid search with 10-fold cross-validation.
- **Prompting:** The question is directly provided to `gpt-4o-mini` without retrieval. The model must infer both the likelihood of disease and its justification to synthesize the encounter note directly from EHR data.
- **RAG:** The question is augmented with the top-5 most similar encounter notes (***k*** = 5) retrieved from the training set using cosine similarity over Clinical ModernBERT embeddings. The retrieved encounter notes are concatenated with the question as supporting evidence from past cases evidence (Figure 1) for generation using `gpt_4o_mini` (temperature = 0). This setup follows standard practice in RAG-based QA configurations (Lewis et al., 2020; Asai et al., 2024)

**Task 1: Binary Decision**

The first task evaluates whether the models can correctly identify patients at high risk of developing csPC and requiring follow-up. Each model outputs a likelihood representing a disease risk estimation. The provided likelihood is then converted into a binary decision label using a fixed threshold and further incorporated into the generated encounter note after thresholding. This setup mirrors how a GP interprets likelihood against risk thresholds when deciding on further follow as depicted in Figure 1.

For consistency, all models produce a likelihood during inference: for KNN, the likelihood corresponds to the fraction of most similar patients with a positive binary decision; for the LLM-based approaches (Prompting and RAG), it is derived from the model’s predicted likelihood or explicit numerical estimate of disease risk. For evaluation purposes, in all cases the threshold was set to 0.5.

**Task 2: Encounter Note Synthesis**

The second task evaluates the models’ ability to synthesize encounter notes that emulate the notes a GP writes after consultation. In the Prompting scenario, the LLM synthesizes a full encounter note directly from the given question, integrating both the likelihood statement and its justification. In contrast, the encounter note synthesis in RAG is conditioned on the retrieved encounter notes from the training set, which serve as a Supporting Evidence.

### Evaluation

Model performance is assessed using both automatic and manual evaluation by a clinical expert with over 5 years of clinical practice experience. The automatic evaluation consists of the binary classification task for predicting whether the patient is likely to develop csPC and thus requires follow-up. The classification is evaluated using balanced accuracy, F1, specificity, sensitivity, and AUC metrics to quantify the model’s discrimination ability between patients requiring follow-up and does that do not (Fernandez-Quilez et al., 2024).

To evaluate the quality of the encounter notes and whether the supporting cases are relevant and similar to the given questions, the manual evaluation is conducted on a subset of 25 cases from the test by an clinical expert. The clinical expert reviews the retrieved supporting cases and the model-predicted encounter notes, answering a structured of Yes/No questions that assess whether the followup recommendation is appropriate, the likelihood estimate clinically reasonable, the justification is clear and whether the supporting retrieved cases are relevant. This evaluation complements the automatic binary decision evaluation and clinical note quality by verifying the human-based clinical plausibility and guideline alignment of the predicted notes. Each synthesized encounter note is also compared with its corresponding ground truth encounter note from the test set using BLEU and ROUGE scores, which capture surface-level textual overlap but not deeper clinical correctness or judgment quality. Results from applying these metrics are depicted in Table 5.

All experiments employ the same stratified train/test split described in Table 1, ensuring that the original dataset distribution of decision labels (i.e. 0 and 1 binary decisions) are preserved across train and test sets.

## 6. Results and Discussion

As shown in Table 4, the KNN baseline performs only above random (Bal. Acc. = 0.57, AUC = 0.6), suggesting that the age, PSA, PSAd and prostate volume features alone cannot clearly separate patient risk levels without additional context (e.g., most similar encounter notes). In contrast, Prompting achieves the best performance (Bal. Acc. = 0.68, AUC = 0.69), indicating that the LLM likely leverages medical information captured during pre-training. RAG reaches similar balanced accuracy but shows higher sensitivity (0.87) and lower specificity (0.29) implying that retrieval increases the model’s ability to detect positive binary decisions but at the cost of introducing more false positives. Although the scores are modest, they are comparable to art results on this collection and reflect the difficulty of early-stage PCa follow-up decision-making (Fernandez-Quilez et al., 2024).

**Table 3:** Examples of encounter notes generated by Prompting and RAG models at the inference stage.

| Method | Example |
| --- | --- |
| Prompting | The patient's age and elevated PSA level suggest a higher risk for prostate cancer. A PSA of 9.3 ng/mL is above the typical threshold for concern, and while the PSAd of 0.25 ng/mL/mL is moderate, the prostate volume is normal. These factors indicate a significant possibility of clinically relevant prostate cancer, but further diagnostic evaluation is needed for a definitive determination. |
| RAG | The patient is 69 years old with a PSA level of 9.3 ng/mL and a PSAd of 0.25 ng/mL/mL, both of which are elevated. Comparing to Case 5 (69 years, PSA 17.8 ng/mL, PSAd 0.23 ng/mL/mL) and Case 1 (PSA 9.4 ng/mL, PSAd 0.25 ng/mL/mL), the patient has similar elevated PSA and PSAd values, which suggest a higher risk for clinically significant prostate cancer. The prostate volume of 37.0 mL is also within the range of concern, supporting the need for further assessment of significant pathology. |

**Table 4:** Comparison of the models on the binary classification of csPA on the PI-CAI QA dataset. Metrics reported include Balanced Accuracy (Bal. Acc.), F1-score (F1), Specificity (Spec.), Sensitivity (Sens.), and Area Under the ROC Curve (AUC).

| Method | Bal. Acc. | F1 | Spec. | Sens. | AUC |
| --- | --- | --- | --- | --- | --- |
| KNN | 0.57 | 0.58 | 0.54 | 0.60 | 0.63 |
| KNN + Encounter Notes Embeddings | 0.57 | 0.56 | 0.58 | 0.55 | 0.56 |
| Prompting | 0.68 | 0.69 | 0.63 | 0.73 | 0.69 |
| RAG | 0.58 | 0.67 | 0.29 | 0.87 | 0.68 |

**Table 5:** Comparison of explanations and ground truth n-gram overlaps based on BLEU and ROUGE (ROUGE-1, ROUGE-2, ROUGE-L) scores for the LLM-based methods.

| Method | BLEU | ROUGE-1 | ROUGE-2 | ROUGE-L |
| --- | --- | --- | --- | --- |
| Prompting | 0.09 | 0.50 | 0.26 | 0.34 |
| RAG | 0.12 | 0.58 | 0.31 | 0.39 |

Table 3 shows examples of predicted encounter notes synthesized with Prompting and RAG settings to compare their generated clinical justifications and follow-up recommendations. During the manual evaluation by the clinical expert, all followup recommendations (% 100) in the tested subset are judged as clinically appropriate by the expert, meaning that the model’s advice for further tests or monitoring matched with PSA and PSAd thresholds in the guidelines. On the other hand, the estimated likelihood suggested by the system is deemed correct in only about half of the cases (% 52). Around half of the justifications (% 52) are also considered clinically clear and aligned with clinical expectations, while most retrieved supporting cases (i.e. similar encounter notes) are assessed as relevant and helpful for synthesizing new patient encounter notes (% 92).

When looking at the retrieved encounter notes in more detail, 100 out of 125 individual retrieved cases (i.e. 5 cases retrieved for each of 25 manually evaluated notes) are marked as useful. After close inspection, we observe that the most of the incorrectly retrieved cases contain big differences between retrieved PSA and PSAd values when compared to those of the patient under consideration. These differences reveal models’ sensitivity to small variations in patient features, which must be further analyzed to understand what patterns the model relies on and how retrieval affects its judgments. Overall, the results show that even though the reliability of the encounter notes is still limited, the RAG is able to quite reliably provide the supporting cases, what might be crucial for subsequent GP’s recommendation.

## 7. Conclusion and Future Work

In this work, we introduce **TabMedQA**, a framework for constructing medical QA collections directly from structured EHRs. By combining instruction-tuned LLMs with disease-specific clinical guidelines, the framework enables the automated generation of clinical encounter notes that simulate how GPs formulate and document early-stage clinical decisions, including the clinically-grounded justifications that support them.

We demonstrate the applicability of **TabMedQA** in a prostate cancer case study, where the synthesized encounter notes include both guideline-based justifications and follow-up recommendations. Experiments show that LLM-based methods make more clinically accurate decisions than feature-based baselines, even though the justifications are not fully clinically correct, factual, or clear. On the other hand, manual expert evaluations show that RAG can reliably retrieve supporting cases and thus enables more reliable human machine-aided decision making.

Future work will extend the framework to additional diseases and multi-modal EHR data, explore automatic factuality and guideline-adherence metrics, and investigate temporally aware RAG setups where retrieved clinical evidence evolves over time.

## 8. Limitations

Despite of the effectiveness of our TabMedQA framework, certain limitations still remain. First, since the questions are derived from structured EHRs rather than real GP-patient interactions, the linguistic diversity of the generated text remain limited. Even though an LLM is employed to enrich phrasing and emulate human questions. As a consequence, the resulting language may still reflect the structured nature of EHRs rather than genuine GP questions. In the same direction, the quality and coherence of the synthesized encounter notes are also dependent on the specific LLM being used during the collection construction, which may introduce model biases or inconsistencies. While our framework leverages medical guidelines, its reliability still depends on expert oversight to ensure that the synthesized justification and follow-up recommendations are truly guidelines-consistent.

Finally, TabMedQA is currently demonstrated on a single structured collection (PI-CAI), which limits its generalization across other clinical domains and data sources.

## Data Availability

All data produced are available on the GitHub repository linked in the article

https://github.com/iai-group/cl4health-tabmedqa/

## 9. Ethics Statements

This research study was conducted using retrospectively open-access human subject data (Saha et al., 2024). The use was approved by the institutional review board of all PI-CAI centers.

## Footnotes

1 https://github.com/iai-group/cl4health-tabmedqa/

2 https://pi-cai.grand-challenge.org/

3 https://pi-cai.grand-challenge.org/DATA/

4 https://github.com/DIAGNijmegen/picai_labels

5 GPT 4o mini Cost Efficient

6 Our adapted guidelines are available at https://anonymous.4open.science/r/lrec2026-tabmedqa-40B9.

7 https://huggingface.co/Simonlee711/Clinical_ModernBERT

